# Trait responsiveness to verbal suggestions predicts placebo responses: A multi-level meta-analysis

**DOI:** 10.64898/2026.08.20.26360892

**Authors:** Madeline V. Stein, Trevor Thompson, Devin B. Terhune

**Author notes:** **Correspondence address:** Madeline V. Stein Department of Psychology, Institute of Psychiatry, Psychology C Neuroscience King’s College London, 16 De Crespigny Park London SE5 8AB, UK. For the purposes of open access, the author has applied a Creative Commons Attribution (CC BY) license to any Accepted Author manuscript version arising from this submission.

## Abstract

**Background:** Placebo responding involves the reduction of symptoms in response to contextual features of an intervention (e.g., verbal suggestions), yet it is characterized by pronounced heterogeneity. Although verbal suggestions are widely recognised as a hallmark method for inducing placebo responses, an open question is whether variability in placebo responding can be partly attributed to individual differences in trait responsiveness to verbal suggestions (REVS). We conducted a pre-registered meta-analysis (PROSPERO registration number CRD420250654692) to quantitatively synthesize available research on the association between trait REVS and placebo responding.

**Methods:** PsycInfo, PubMed, MEDLINE, and Embase were searched up to June 2026 for original clinical or experimental studies involving both the assessment of REVS and symptom measures (self-report, behavioural, and/or physiological) in response to an inactive intervention (placebo).

**Results:** Of 1,512 search results, 24 articles presenting 66 correlations between REVS and placebo responding were analysed (*N* = 1,137). A multi-level meta-analysis revealed a significant, albeit weak, positive correlation between REVS and placebo responses, *r* = 0.18 [95% CI: 0.13, 0.24], such that individuals with higher REVS reported greater symptom relief in response to the placebo. Meta-regression analyses did not identify any significant moderators of the correlation between REVS and placebo responding and sensitivity analyses based on Bayesian subgroup estimates indicated that the aggregate correlation was stable across methodological quality indicators and study features.

**Conclusion:** These findings suggest that individual differences in REVS may partly explain variability in symptom reduction in response to placebos, with implications for the sources of variance in placebo effects in experimental and applied contexts.

## Introduction

Placebo responses highlight how communication, expectations, learning, and social context can profoundly shape perception, physiology, and clinical outcomes [1]. They play a paramount role in patients’ treatment responses across medical and mental health symptoms, influencing how patients respond to both inert interventions [2] and active treatments [3]. Placebo responding shows substantial interindividual variability, such that some individuals experience marked symptom improvement whereas others do not [4]. The reliable identification of putative good placebo responders has salient implications spanning both basic science and clinical application: identifying such individuals could help to minimize placebo responses in treatment trials (thereby enhancing internal validity), augment placebo responsiveness to boost treatment outcomes, and facilitate precision medicine approaches involving tailored treatment planning based on an individual’s neurocognitive profile [5]. Nevertheless, evidence that placebo responding is stable across contexts is inconsistent [6–8], and whether placebo responding constitutes a stable trait-like variable remains controversial [9]. Consistent with this uncertainty, attempts to identify reliable trait markers of placebo responsiveness have largely been unsuccessful [10, 11].

One possible explanation for the difficulty in identifying reliable predictors of placebo responding is that previous meta-analyses have not focused on individual difference variables that moderate response to placebo induction procedures, which may represent more promising candidate predictors. Among established methods for reliably eliciting placebo responses [12, 13, 11], verbal suggestions are widely recognised as central to the reduction of symptoms across clinical and experimental contexts [2], and are arguably the most easily harnessed within clinical encounters [14]. Suggestions are communications capable of producing and modifying psychological and physiological processes that are presented in a way that the response will be experienced as something that *happens* to the individual rather than as a deliberate action [15, 16]. Across contexts, suggestions are often embedded within communications regarding an intervention or procedure either directly (e.g., “this procedure will decrease your pain”) or indirectly (e.g., “some people report feeling less pain”) [17–19]. Meta-analytic evidence has shown that verbal suggestions alone can induce small-to-medium placebo responses across a variety of symptom domains [12, 13, 20].

The pronounced role of suggestion in the induction of placebo responses indicates that trait responsiveness to verbal suggestions (REVS; [21]) could be a valuable starting point to elucidate individual differences in placebo responding [21]. REVS is highly stable across the lifespan [22], and follows a normal distribution in the general population (e.g., [23, 19]). It is a reliable predictor of responsiveness to hypnotic hypoalgesia suggestions in clinical [24] and experimental contexts [25]. It is also a reliable, albeit weak, predictor of nocebo responding [26], a phenomenon that is related to, but distinct from, placebo responding [27]. Multiple experimental studies have reported positive associations between REVS and placebo responding [28, 29] although this association has been somewhat inconsistent across studies [30, 31]. These inconsistencies are plausibly attributable to methodological differences, such as the mode (e.g., verbal) or type of suggestion (e.g., direct), the concurrent use of conditioning, the target symptom (e.g., pain), and/or participant expectations about potential symptom relief [28]. Previous reviews and meta-analyses have largely overlooked the potential role of REVS in placebo responding and, to our knowledge, the magnitude of this association has not been formally quantified.

This meta-analysis sought to quantitatively synthesize the extant literature in order to estimate the magnitude of the association between REVS and placebo responding. Toward this end, we conducted a pre-registered (PROSPERO registration number: CRD420250654692) multi-level meta-analysis in accordance with Meta-analyses of Observational Studies in Epidemiology (MOOSE) [32] and Preferred Reporting Items for Systematic Reviews and Meta-Analyses (PRISMA) guidelines [33]. We systematically integrated research studies examining the correlation between standardized REVS measures and symptom outcomes in response to an inactive (sham) intervention. In order to clarify sources of variability in this association, we also examined whether these correlations were moderated by the corresponding magnitudes of expectancy and placebo effects, as well as other methodological features.

## Methods

### Eligibility criteria

Our pre-registered inclusion criteria were: published in English; published in an academic peer-reviewed journal after 1958, corresponding to the introduction of the first standardized hypnotic suggestibility scale [34]; use of one or more communicative (e.g., verbal suggestion) or learning (e.g., conditioning) manipulations to promote a placebo response (i.e., change in a target symptom or outcome); administration of an inactive (sham) intervention; inclusion of a symptom measure in response to the placebo intervention; and inclusion of a standardized REVS (suggestibility) measure. We employed broad inclusion criteria to obtain an accurate representation of the REVS-placebo responding literature: this included studies in which the placebo intervention was administered within a hypnotic context, outcomes not uniformly experienced as pleasant (e.g., intoxication), and studies without a control condition, given that the absence of a comparator condition was anticipated to be a common limitation.

### Search strategy

PsycInfo, PubMed, MEDLINE, and Embase were searched in February 2025; the search was repeated until June 2026 (yielding one new article). The search string ((placebo* OR nocebo*) AND (suggestib* OR hypnosis OR hypnotizab* OR hypnotisab* OR “hypnotic susceptib*”)) was augmented with reference list screening of all included articles and relevant reviews.

### Study selection

Three reviewers independently screened titles and abstracts of articles returned by initial searches; articles that did not meet our eligibility criteria were rejected. The full texts of the remaining articles were independently reviewed by the same three reviewers, who then compiled a final list of articles. Disagreements at either stage were resolved through discussion with the authors (MVS, TT, and DBT). If a paper met our inclusion criteria but did not report the relevant statistics and was published ≥ 10 years ago, it was excluded as we anticipated the data would not be accessible, resulting in the exclusion of seven papers. Seven author groups were contacted to provide missing data; of these, three no longer had access to necessary data, resulting in exclusion, whereas the remaining four provided sufficient information to permit inclusion. When relevant data were presented in a figure (two papers), WebPlotDigitizer (version 4.6; [35]) was used to extract relevant outcome data from the figure in lieu of requesting the data from the authors.

### Data extraction

Full-text data extraction was performed independently by two reviewers. The primary outcomes were correlation coefficients between a REVS scale and placebo response (operationalised as the respective outcome measure in the placebo condition alone [no control], placebo-control, or placebo-baseline). Studies were coded for the following criteria: study information (authors, publication year, journal, title, country); demographic information (sex assigned at birth distributions, age, ethnicity, education); sample (e.g., clinical, non-clinical, all/some students); study design (within-groups, between-groups); symptom domain (e.g., pain, itch, dyspnoea, dizziness, nausea, motor inhibition, cognitive performance, affect, skin condition); methodological details (counterbalancing of placebo administration and REVS scale measurement, participant and experimenter blinding of condition [yes vs. no]; experimenter blinded to REVS and/or placebo response magnitude when assessing the other [yes vs. no]; randomisation to allocated condition [yes vs. no]; experimenter sex assigned as birth; sample size [total and for all conditions/groups]; expectancy assessed prior to placebo administration [yes vs. no]; and expectancy descriptive statistics [*Ms*, *SDs*] in placebo and control conditions; placebo suggestion modality [absence/presence of: verbal suggestion, textual suggestion, non-verbal suggestion]; suggestion type [direct or indirect]; inert intervention exposure [pill, sham device [e.g., sham wi-fi exposure], nasal spray, cream, injection, inhalant, etc.]; use of conditioning [yes vs. no]; use of social observation [yes vs. no]; placebo information type [open vs. deceptive]; symptom score descriptive statistics [*M*s, *SD*s] in placebo and control conditions; REVS scale administered in hypnotic context [yes vs. no]; scale type [direct, indirect, retrospective]; REVS scale administration context [group vs. individual]; and good psychometric properties for the REVS scale according to relevant reliability and validity data presented in the paper [yes vs. no]). The reviewers displayed acceptable agreement (84%), and all extracted data were re-reviewed by MVS and DBT, with any discrepancies resolved through discussion.

### Study methodological quality

A 15-item scale was developed to assess study methodological quality (see **Supplementary Methods**). Items were adapted from a previous measure [25, 36, 37, 26, 38] and based on Cochrane criteria [39] and PRISMA [33] recommendations. The two reviewers independently rated each item categorically (0 = criterion not met, 1 = criterion met); when information pertaining to a criterion was not reported, or could not be reasonably inferred, the respective criterion score was rated as not met, and a total score (percentage of criteria met) was computed for each study. Agreement between reviewers was fair (percentage agreement: 74%; Cohen’s kappa =.49) and all discrepancies were resolved through discussion with MVS and DBT.

### Data synthesis

All analyses were completed in *R* [40]. Individual study effect sizes included unadjusted correlation coefficients between REVS scale scores and placebo responses that were transformed to *z*-scores using Fisher’s *r*-to-*z* transformation. We report back-transformed correlation coefficients (*r_bt_*) for ease of interpretation, with positive values representing a positive association between REVS and placebo response. Individual study effect sizes for placebo responses and placebo expectancy effects were calculated as within-group differences between placebo and control conditions (when available), using Hedges’ *g* as the standardized measure of effect size. When necessary, *SD*s were calculated from standard errors or 95% confidence intervals (CIs). Positive values reflect larger placebo responses and expectancy effects.

Random-effects and multi-level meta-analyses were performed using the metafor package [41]. As individual studies could contribute more than one effect size (e.g., across repeated time assessments, multiple outcomes), a three-level meta-analytic model was employed to account for the hierarchical structure of the data, partitioning variance into three levels: (1) sampling variance for individual effect sizes, (2) within-study variability among multiple effect sizes reported within the same study, and (3) between-study variability across studies. Multi-level analyses were conducted by comparing the three-level model to a reduced two-level model (in which the effect size–level random effect was removed) using the Akaike Information Criterion (AIC), Bayesian Information Criterion (BIC), and a likelihood ratio test (LRT; [42]. For all tested models, the three-level model had lower AIC and BIC values and a significant LRT, reflecting superior fit. No studies were identified as outliers on the basis of studentized residuals (|*Z*|>3) within the main analysis sample (REVS-placebo response), or symptom expectancy and placebo effect subgroup analyses [43]. Heterogeneity was computed through Cochran’s *Q*, *I*^2^ and *τ*^2^, where larger *Q* values reflect greater variability in effect sizes beyond that expected by sampling error, *I*^2^ ≥ 50% indicates moderate or greater heterogeneity, and τ^2^ describes the variance of true effect sizes. Jackknife analyses, in which each correlation pair was sequentially omitted and the analysis was reperformed, were conducted to evaluate the robustness of the pooled association by assessing the influence of individual studies on the overall effect estimate [43]

We conducted Egger’s regression tests [44] to assess potential publication bias in a random-effects model, adopting a threshold of *p* < .10 to reduce the risk of Type II errors [45] and visually inspected funnel plots of effect sizes against their standard errors. We also estimated asymmetry-corrected cumulative effect sizes using the trim-and-fill method [46]. In addition, we ran an Egger-style meta-regression within the multi-level model by including study standard error as a moderator, allowing us to evaluate potential small-study effects while accounting for clustering of effect sizes within studies [47].

Meta-regression analyses were undertaken to account for heterogeneity in effect sizes. The corresponding statistic (*Z_b_*) represents the correlation difference between the two levels of the moderator on the Fisher’s *z*–transformed correlation scale and was not back-transformed to the correlation metric. Categorical moderator variables with > 2 levels were decomposed into simpler two-level moderators. Moderation analyses were undertaken when ≥ 2 studies per level of the moderator were available from at least two independent studies. Moderators included 35 categorical variables: target symptom domain (pain, cognitive function, affect); placebo induction method (verbal, textual, non-verbal, conditioning); suggestion type (0 = indirect, 1 = direct); placebo administration procedure (pill, cream, sham device, inhalant); features of REVS measurement (group vs. individual administration, hypnotic context, assessed in same context as placebo administration); application of counterbalancing; assessment of expectancy (yes vs. no); experimenter sex (female vs. male, mix of male/female vs. sole female experimenter); sample type (clinical vs. non-clinical, some/all students v. non-student); and three continuous variables: methodological quality (percentage score), and the magnitudes of the expectancy and placebo effect. Pre-specified and exploratory subgroup analyses were conducted as stratified meta-analyses within moderator-defined subsets (≥5 effect sizes from ≥2 studies).

To assess whether primary and subgroup effect sizes were small enough to be considered practically equivalent to zero, we deviated from our pre-registration (due to the multi-level structure of the data) and conducted Bayesian equivalence analyses using a Region of Practical Equivalence (ROPE) approach [48]. We applied a predefined equivalence margin of ± 0.10 on Fisher *z*-transformed correlation coefficients, consistent with small effect sizes [49]. Evidence for practical equivalence was evaluated by examining the proportion of the posterior distribution falling within the ROPE (i.e., equivalence margin), with effects considered non-negligible when the posterior mass within the ROPE was small [50]. In particular, small proportions of posterior mass (≤ 10%) within the ROPE were taken as evidence that the subgroup-specific effect was meaningfully different from zero, whereas large proportions (≥ 30%) were interpreted as reflecting practical equivalence, and intermediate values (≈10–30%) were interpreted as ambiguous [48]. These ROPE assessments were used to characterise the magnitude and practical relevance of subgroup differences and moderation effects.

## Results

A PRISMA diagram presenting study selection can be found in **Figure 1**. The final sample of 24 included papers reported 26 studies from which 66 REVS-placebo response correlation coefficients were extracted, with 17 papers reporting more than one correlation (*M* = 2.75, *SD* **=** 1.80; range = 1 – 8). Details of the included studies can be found in **Table 1**, and a full list of included papers can be found in the **Supplementary Results**.

**Figure 1.**
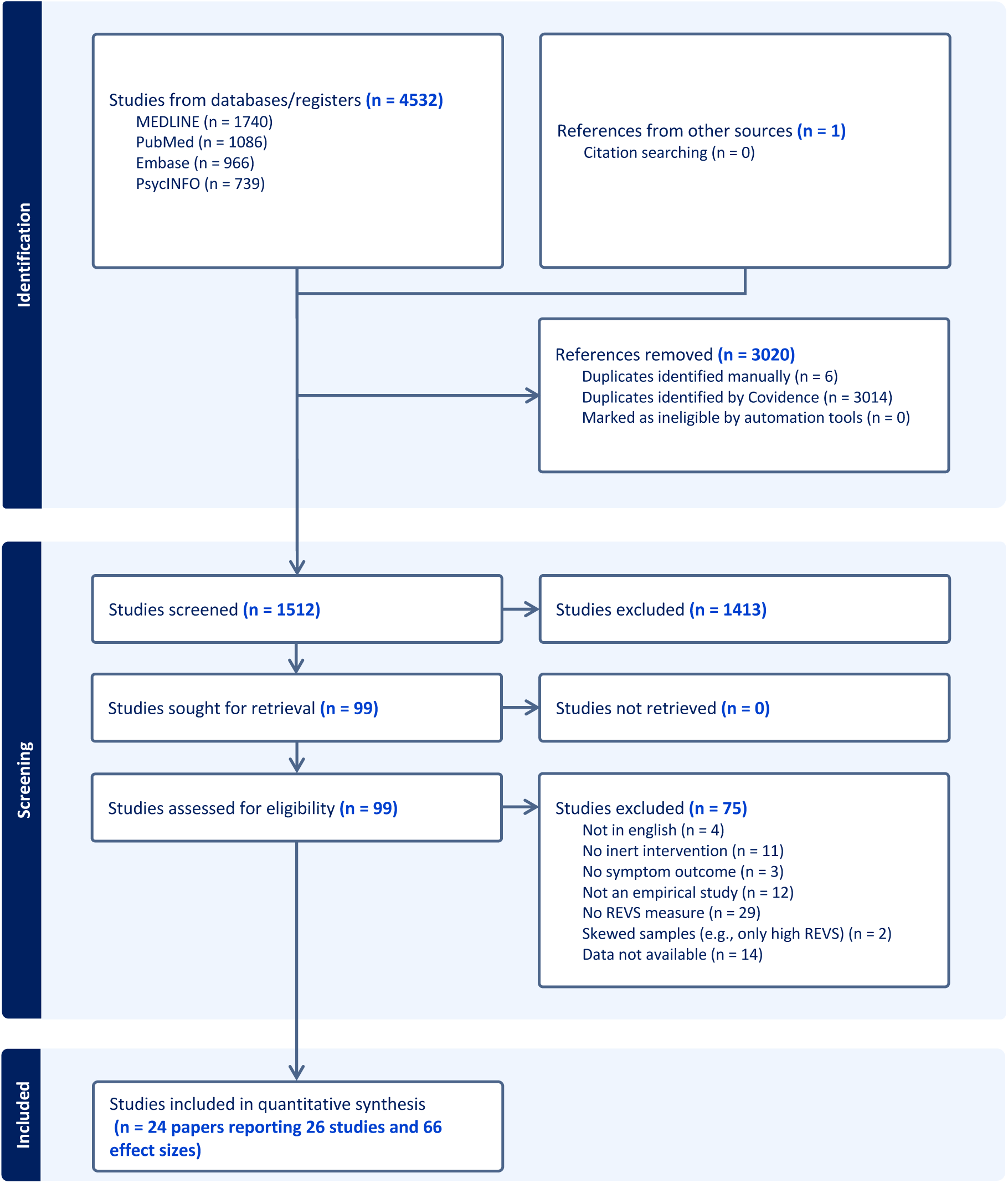
PRISMA flowchart of the study selection process.

**Table 1.** Principal features of included studies reporting correlations between REVS and placebo responses (n = 2C).

| Source | # of correlations | Sample | N (% female) | Symptom domain(s) | Symptom outcome measure(s) | Placebo type | Placebo induction method(s) | Reference condition | REVS scale(s) | REVS scale type |
| --- | --- | --- | --- | --- | --- | --- | --- | --- | --- | --- |
| Baker et al., 1993* | 4 | Some/all psychology students | 69 (62%) | Pain | VAS | Inhalant | Verbal suggestion | Baseline | HGSHS:A | Direct |
| Bentler et al., 1963 | 2 | Female nursing students | 19 (100%) | Pain | Threshold; tolerance | Pill | Verbal suggestion | Control | SHSS:A | Direct |
| Bush & Ditto 1985 | 1 | Chronic back pain patients | 22(-) | Pain | Index composite change score | Sham-device | Textual suggestion | - | SHALIT | Direct |
| Corsi & Colloca 2017 | 5 | - | 46 (52%) | Pain | VAS | Procedure | Conditioning | Control | MISS | Retrospective |
| De Pascalis & Scacchia, 2019 | 4 | Women volunteers | 58 (100%) | Pain; affect | NRS pain; NRS distress; HRV | Cream | Verbal suggestion | Baseline | SHSS:C | Direct |
| De Pascalis et al., 1999 | 4 | Right-handed non-clinical controls | 29 (100%) | Pain; affect | Pain rating; distress rating; SCR; Pain thresholds; Sensory thresholds | Cream | Verbal suggestion | Baseline | SHCS | Direct |
| De Pascalis et al., 2001 | 3 | Some/all undergraduate students | 29 (100%) | Pain | Pain rating; distress rating; SCR | Nasal spray | Textual suggestion | Control | SHSS:C | Direct |
| De Pascalis et al., 2002 | 4 | Some/all students | 36 (-) | Pain; affect | Pain intensity; Pain unpleasantness | Cream | Verbal suggestion | Control | SSS | Indirect |
| De Pascalis et al., 2016* | 4 | Some/all students | 36 (100%) | Pain; affect | NRS pain; NRS distress | Cream | Verbal suggestion | Baseline | Italian SHSS:C | Direct |
| De Pascalis et al., 2021* | 2 | Right-handed women | 56 (100%) | Pain | NPS | Cream | Verbal suggestion | Control | SHCS | Direct |
| Guestini & Parris, 2025 | 2 | Some/all students | 48 (54%) | Pain | Pain threshold; Pain tolerance | Cream | Verbal suggestion | Control | Modified-SHSS:C | Direct |
| Huber et al., 2013 | 1 | - | 28 (60%) | Pain | VAS | Sham-device | Verbal suggestion; conditioning | - | Italian-SHSC:A | Direct |
| Leigh et al., 2003 | 1 | Asthma patients | 17 (64%) | Dyspnoea | Borg dyspnoea scale | Inhalant | Verbal suggestion | - | CIS | Direct |
| Lifshitz et al., 2017 | 5 | Some/all psychology students | 15 (-) | Affect | Likert relaxation; Likert drowsiness; Systolic BP; Diastolic BP; HR | Pill | Verbal, non-verbal suggestions | Baseline | HGSHS:A | Direct |
| Lund et al., 2015 | 2 | Some/all students | 44 (-) | Pain | Pain intensity; Peak pain | Injection | Verbal suggestion; conditioning | Control | SSG | Indirect |
| McGlashan et al., 1969 | 1 | Some/all students | 24 (0%) | Pain | Total tolerance | Pill | Verbal suggestion | Baseline | HGSHS:A, SHSS:C | Direct |
| Parsons et al., 2021-Study 1 | 1 | Some/all psychology students | 57 (77%) | Pain | VAS | Cream | Verbal suggestion; conditioning | Control | MHGS | Direct |
| Parsons et al., 2021-Study 2 | 1 | Some/all students | 78 (65%) | Pain | VAS | Cream | Verbal suggestion | Control | BSS | Direct |
| Ryan & Souheaver, 1976 | 1 | Psychiatric inpatients | 12 (-) | Affect | STAI | Sham-device | Verbal suggestion | Baseline | HGSHS:A | Direct |
| Sharav et al., 2023 | 3 | Non-clinical controls | 24 (62%) | Pain | VAS | Cream | Verbal suggestion; conditioning | Baseline | SHALIT | Abbreviated direct |
| Sheiner et al., 2016 | 5 | Some/all psychology students | 22 (-) | Affect | Likert relaxation; Likert drowsiness; Systolic BP; Diastolic BP; HR | Pill | Verbal, textual, non-verbal suggestion; | Baseline | HGSHS:A | Direct |
| Spanos et al., 1988* | 2 | Some/all students with hand warts | 24 (-) | Skin condition | Wart decrease | Sham-device | Verbal, textual, non-verbal suggestion; | Baseline | CURSS | Direct |
| Spanos et al., 1989- Study 1* | 1 | Some/all undergraduate students | 64 (-) | Pain | Intensity rating | Cream | Verbal suggestion | Baseline | CURSS | Direct |
| Spanos et al., 1989- Study 2* | 4 | Some/all undergraduate students | 60 (-) | Pain | Intensity rating | Cream | Verbal suggestion | Baseline | CURSS | Direct |
| Tasso et al., 2020 | 1 | Some/all psychology students | 110 (70%) | Cognition; affect | Placebo response | Sound | Verbal, textual, non-verbal suggestions | - | HGSHS:A | Direct |
| Woody et al., 1997 | 2 | Some/all undergraduate students | 93 (-) | Dizziness; nausea; motor performance; cognition | Subjective experience measure; Unsuggested symptom checklist | Drink | Verbal, non-verbal suggestions | - | HGSHS:A | Direct |

**Notes.** -, Not reported; \*, correlations where placebo suggestions were administered in a hypnotic context; VAS, Visual Analogue Scale; NPS, numerical pain scale; STAI, State Anxiety Inventory; SCR, skin conductance; NRS, numerical rating scale; BP, blood pressure; HR, heart rate; Control comparator relative to extracted correlation used in synthesis; MHGS, modified-Harvard Group Scale of Hypnotic Susceptibility; BSS, Brief Suggestibility Scale; HGSHS:A, Harvard Group Scale of Hypnotic Susceptibility: Form A; MISS, Multidimensional Iowa Suggestibility Scale; SHSS:C, Stanford Hypnotic Susceptibility Scale: Form C; SSS, Sensory suggestibility Scale; SHCS, Stanford Hypnotic Clinical Scale; CURSS, Carleton University Responsiveness to Suggestion Scale; SHALIT, Six-minute Arm Levitation Test; CIS, Creative Imagination Scale.

### Methodological quality criteria

Methodological quality scores were relatively low across the 26 included studies, with most studies meeting fewer than half of the assessed criteria (*M* = 48%, *SD* = 17%; range: 14–78%; see **Supplementary Table 1**). All studies clearly reported their aims, but only 10% (*n* = 7) reported relevant reliability data for the REVS measure. A minority of studies implemented precautions to ensure that experimenters were blinded to the experimental conditions (34%; *n* = 9), and fewer than half (42%; *n* = 11) ensured blinding of experimenters to participants’ REVS scores when assessing placebo responses. In contrast, 76% (*n* = 20) ensured experimenters were blinded to participants’ placebo responses when measuring REVS and over half (57%; *n* = 15) ensured that participants were blinded to experimental conditions. Finally, with respect to open science practices, only one study (3%) was prospectively pre-registered, and two studies (7%, from the same paper) made their data publicly available on an open repository (e.g., Open Science Framework).

### Meta-analysis of bivariate correlations between REVS and placebo responses

A multi-level meta-analysis of correlation coefficients between symptom outcomes in response to an inactive (sham) procedure and REVS scores (*k* = 66, *n* = 25^1^) revealed a significant association, *r_bt_* = 0.18 [95% CI: 0.13, 0.24], *Z* = 6.73, *p* <.001 (**Figure 2**). 59 of the correlation coefficients (89%) were positive in direction. A Jackknife analysis confirmed the aggregate effect size narrowly varied in the weak range (*r_bt_* range: 0.17 – 0.19) and the cumulative effect remained statistically significant in all 66 iterations, indicating that it was not driven by a single effect size. The magnitude of heterogeneity was low (*Q*_(65)_ = 91.89, *p* = .015), with negligible within-study (*I^2^* = 0%, τ² = 0.00) and low between-study variance (*I^2^* = 15%, τ² = 0.01) indicating that sampling error accounted for most of the observed variability (*I ^2^* = 16%). Finally, Bayesian ROPE analyses indicated that the pooled association was not practically equivalent to zero: the posterior median correlation was *r* = 0.19 [95% CrI: 0.12, 0.24], and only 0.6% of the posterior mass fell within the ROPE (−0.10 ≤ *r* ≤ 0.10), providing strong evidence against practical equivalence to zero. Cumulatively, these results demonstrate that REVS is a significant, albeit weak, predictor of placebo responding.

**Figure 2.**
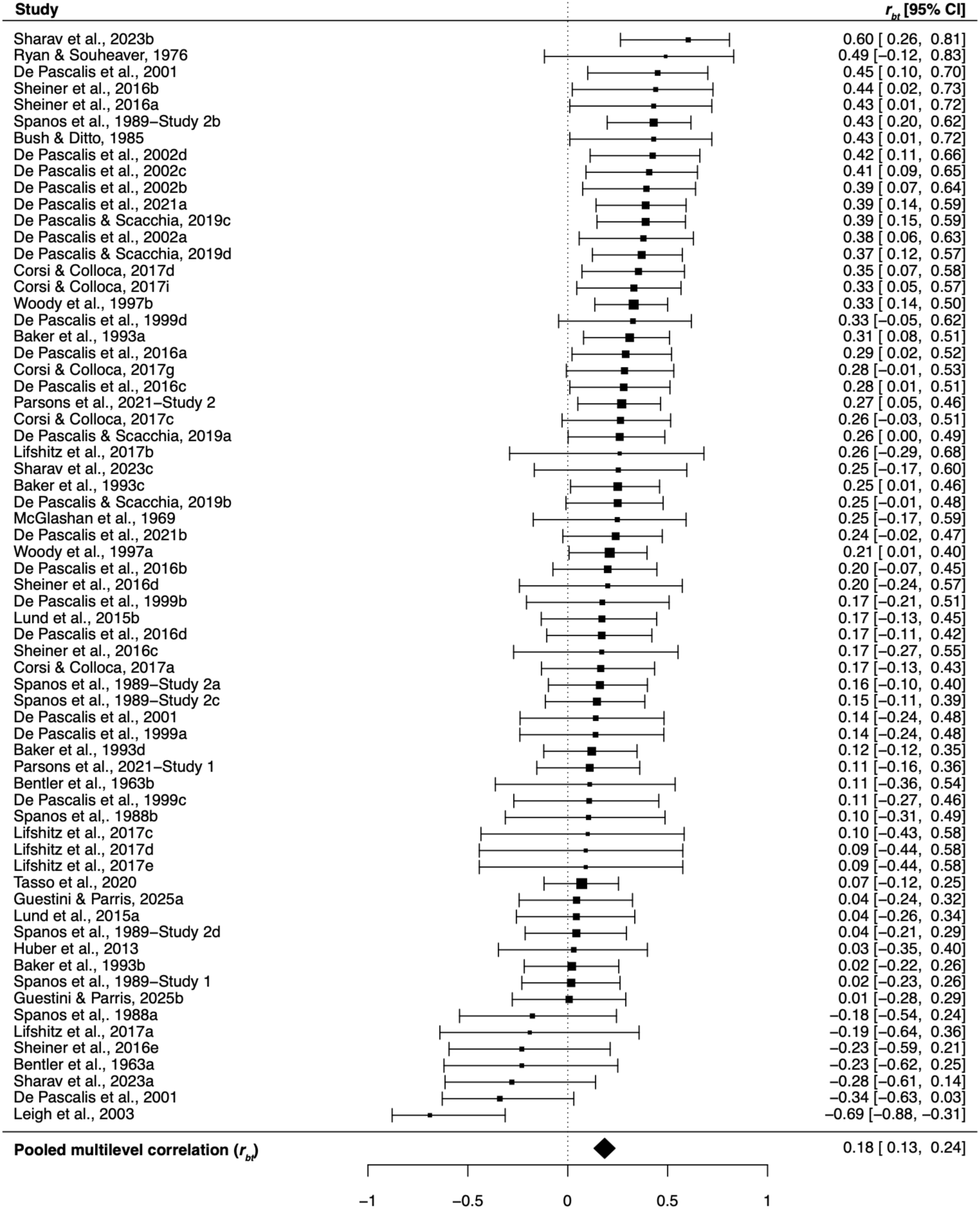
Forest plot of back-transformed REVS-placebo response correlation coefficients (k = CC; n = 25*). **Notes.** Effect sizes were analysed using Zr but plotted here as Pearson’s r (rbt) for ease of interpretation. Marker sizes reflect study weights, with smaller and larger markers denoting smaller and larger weights, respectively. * = Two studies that reported overlapping samples were clustered together for the analyses.

### Publication bias

Egger’s regression test and a funnel plot did not suggest any evidence of asymmetry within a random-effects model (*Z* = –1.56, *p* = .12; **Figure 3**) or a multi-level model of the data (β = –0.76, *SE* = 0.56, *Z*= –1.36, *p* = .18). Indeed, a trim-and-fill analysis did not impute any new effects, and the adjusted pooled estimate, *r_bt_* = 0.20 [0.16, 0.24], was very similar to the original estimate.

**Figure 3.**
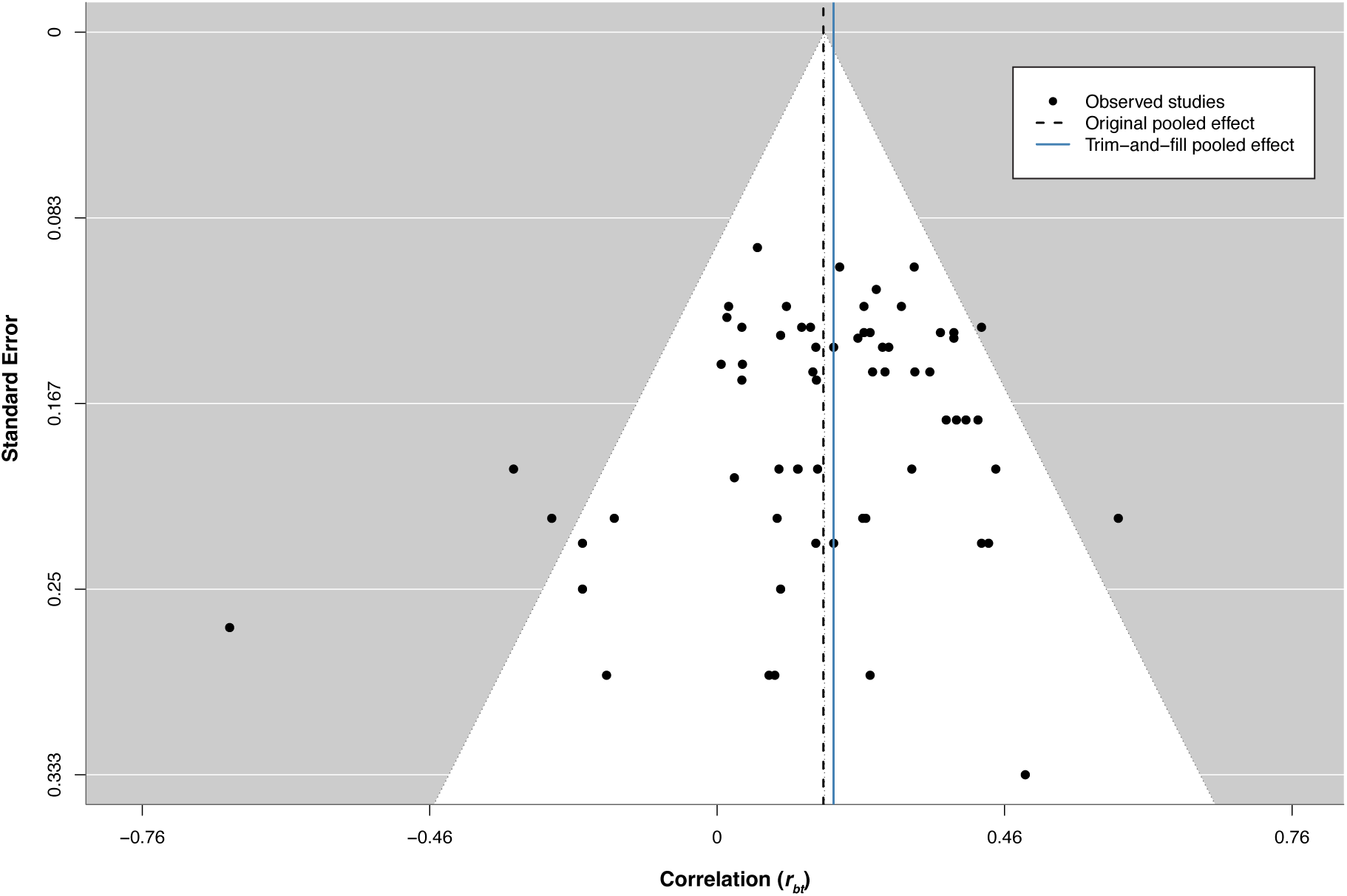
Random-effects funnel plot of REVS-placebo back-transformed correlation coefficients as a function of standard errors (k = CC). **Notes.** Markers denote individual study effect sizes and standard errors.

### Meta-analysis and meta-regression of placebo expectancy and placebo effects

A random-effects meta-analysis of studies assessing symptom reduction expectations for the placebo procedure (*k* = 10) indicated a large overall placebo expectancy effect, *g* = 1.97 [1.07, 2.83], Z = 4.36, *p* < .001, albeit with considerable heterogeneity, *Q*_(9)_= 187.94, *p* <.001, *I^2^* = 97%, τ² = 1.93 (**Supplementary Figure 1**). Conversely, the overall magnitude of the placebo effect within a multi-level meta-analysis was weak, albeit significant (*k* = 15, *n* = 9), *g =* 0.37 [0.12, 0.63], Z = 2.93, *p* = .003, with moderate heterogeneity (*Q*_(14)_= 82.54, *p* <.001) between studies (*I^2^* = 68%, τ² = .12), but not within (*I^2^* = 0%, τ² = .00) (**Supplementary Figure 3**).

Within these two study subgroups, the magnitudes of both the placebo expectancy effect and the placebo effect did not significantly moderate the correlation between REVS and placebo responding (**Table 2**), although in the case of the latter, it showed a non-significant positive trend, reflecting that studies with larger placebo effects tended to exhibit stronger associations between REVS and placebo responding. Relatedly, in the full sample, effect sizes were not significantly larger in studies that assessed expectancy or the placebo effect (i.e., placebo-control) (**Table 2**). However, within a random-effects model, placebo expectancy effects moderated the magnitude of the placebo effect in studies that assessed both expectancy and symptom outcomes in placebo and control conditions (*k* = 5), Δ*g* = 0.46 [0.20, 0.73], Z = 3.47, *p* < .001, with moderate heterogeneity (*Q*_(4)_= 11.98, *p* <.001; *I^2^*= 68%, τ² = .05), indicating that placebo effects tended to be greater with larger placebo expectancy effects.

**Table 2.** Meta-regression analyses on REVS-placebo response correlation coefficients.

| Moderator | Z<sub>b</sub> [95% CIs] | Z | p | B-I<sup>2</sup> | W-I<sup>2</sup> | n (k) |
| --- | --- | --- | --- | --- | --- | --- |
| **Methodological quality** |  |  |  |  |  |  |
| Item 1: relevant reliability data provided for REVS measure | 0.02 [-0.15, 0.21] | 0.30 | .76 | 17% | 0% | 25 (66) |
| Item 2: standardized symptom outcome | 0.04 [-0.07, 0.16] | 0.70 | .47 | 14% | 1% | 25 (66) |
| **Item 4: clear sample recruitment site** | **-0.11 [-0.21, -0.00]** | **-2.08** | **.036** | **1%** | **1%** | **25 (66)** |
| **Item 5: clear participant recruitment strategy** | **-0.09 [-0.18, 0.00]** | **-2.09** | **.035** | **8%** | **1%** | **25 (66)** |
| **Item 6: clear inclusion & exclusion criteria** | **0.10 [0.01, 0.19]** | **2.38** | **.016** | **8%** | **1%** | **25 (66)** |
| Item 7: clear protocol reporting | 0.03 [-0.06, 0.13] | 0.70 | .47 | 13% | 3% | 25 (66) |
| **Item 8: experimenter blind to placebo response when measuring REVS** | **0.17 [0.03, 0.29]** | **2.50** | **.012** | **3%** | **4%** | **25 (66)** |
| Item 9: experimenter blind to REVS when measuring placebo | -0.10 [-0.20, 0.00] | -1.88 | .059 | 8% | 6% | 25 (66) |
| Item 10: participant condition blinding | 0.08 [-0.00, 0.17] | 1.79 | .072 | 6% | 3% | 25 (66) |
| Item 11: experimenter condition blinding | 0.02 [-0.08, 0.12] | 0.43 | .66 | 14% | 2% | 25 (66) |
| Item 12: adequate description of sample demographics | -0.02 [-0.12, 0.08] | -0.40 | .69 | 17% | 0% | 25 (66) |
| Total methodological quality score | 0.00 [-0.00, 0.00] | 0.36 | .71 | 15% | 1% | 25 (66) |
| **Methodological features** |  |  |  |  |  |  |
| Inclusion of control condition | 0.08 [-0.03, 0.19] | 0.15 | .15 | 6% | 1% | 25 (66) |
| Female experimenter | -0.06 [-0.24, 0.11] | -0.75 | .49 | 12% | 12% | 6 (15) |
| Female & male experimenters | 0.12 [-0.04, 0.29] | 1.40 | .15 | 12% | 12% | 6 (15) |
| Non-clinical sample | 0.15 [-0.17, 0.44] | 0.92 | .35 | 15% | 3% | 25 (62) |
| **Some/all student sample** | **0.43 [0.03, 0.71]** | **2.12** | **.036** | **10%** | **4%** | **19 (46)** |
| Magnitude of the placebo effect | 0.22 [-0.00, 0.43] | 1.91 | .055 | 2% | 1% | 9 (15) |
| Measured expectancy | 0.05 [-0.05, 0.16] | 1.00 | .31 | 14% | 1% | 25 (66) |
| Magnitude of expectancy effect* | 0.02 [-0.05, 0.10] | 0.59 | .54 | 4% | 3% | 7 (10) |
| **Placebo administration type** |  |  |  |  |  |  |
| Placebo pill | -0.06 [-0.23, 0.10] | -0.76 | .44 | 14% | 1% | 25 (66) |
| Placebo sham device | -0.04 [-0.26, 0.18] | -0.37 | .71 | 14% | 1% | 25 (66) |
| Placebo cream | 0.07 [-0.02, 0.18] | 1.43 | .14 | 13% | 1% | 25 (66) |
| Placebo inhalant | -0.13 [-0.36, 0.05] | -1.43 | .15 | 2% | 2% | 25 (66) |
| Use of conditioning procedure | -0.00 [-0.14, 0.13] | -0.01 | .99 | 17% | 0% | 25 (66) |
| **Suggestion mode and type** |  |  |  |  |  |  |
| Textual suggestion | -0.07 [-0.21, 0.06] | -1.07 | .28 | 11% | 1% | 25 (66) |
| Verbal suggestion | -0.06 [-0.23, 0.10] | -0.74 | .45 | 17% | 0% | 25 (66) |
| Nonverbal suggestion | -0.04 [-0.18, 0.08] | -0.69 | .48 | 15% | 1% | 25 (66) |
| Use of direct verbal suggestion | -0.14 [-0.28, 0.00] | -1.90 | .056 | 9% | 4% | 9 (20) |
| **REVS assessment features** |  |  |  |  |  |  |
| Group administration of REVS scale | 0.00 [-0.11, 0.13] | 0.15 | .87 | 1% | 3% | 21 (58) |
| Use of two REVS scales | -0.12 [-0.39, 0.17] | -0.82 | .40 | 13% | 1% | 25 (66) |
| REVS scale administered in a hypnotic context | -0.06 [-0.17, 0.04] | -1.16 | .24 | 12% | 0% | 25 (66) |
| Assessment of REVS in the same context as placebo administration | 0.02 [-0.11, 0.15] | 0.29 | .76 | 17% | 0% | 23 (63) |
| **Target symptom domain** |  |  |  |  |  |  |
| Pain | 0.08 [-0.03, 0.21] | 1.36 | .17 | 15% | 1% | 25 (66) |
| Cognition | 0.00 [-0.17, 0.18] | 0.03 | .96 | 15% | 1% | 25 (66) |
| Affect | 0.02 [-0.08, 0.13] | 0.40 | .68 | 15% | 2% | 25 (66) |

Notes: Z<sub>b</sub> = difference between moderator levels on the Fisher z-transformed correlation scale; all moderators were binary except Total methodological quality, magnitude of expectancy effect and magnitude of placebo effect; item refers to the methodological quality criteria (see Supplementary results); for all binary moderators, 0 = variable absent, 1 = variable present.

Taken together, these findings suggest that while placebo expectancy effect and placebo effect magnitudes are related at the study level, neither robustly moderates the association between REVS and placebo responding, although the placebo effect shows a near-significant positive trend.

### Meta-regression analyses

We next conducted a series of exploratory meta-regression analyses to identify variables that moderated the strength of the aggregate association between REVS and placebo responses. Due to our inclusion criteria of at least two studies per binary moderator level from two unique studies, 11 (of 15) methodological quality items were eligible for analysis and were included (**Table 2**). Among these methodological items, studies that clearly reported their inclusion criteria and blinded experimenters to participants’ placebo responses were weak but positive moderators and had larger REVS-placebo response correlations than studies without these features. Conversely, clear reporting of recruitment site and strategy emerged as weak, negative moderators, such that studies that clearly reported these features exhibited weaker correlations than those that did not. There were no additional significant moderating effects associated with individual methodological criteria or with total methodological quality scores. However, experimenter blinding to participants’ placebo responses during REVS assessment showed a non-significant trend as a weak, negative moderator. Bayesian ROPE analyses indicated that moderation effects were often practically equivalent to zero (**Supplementary Table 2**). In contrast, REVS–placebo correlations within subgroups defined by these methodological features were frequently practically meaningful (**Supplementary Table 3**).

Taken together, these results highlight the importance of rigorous experimenter blinding and transparent reporting practices but broadly demonstrate that the REVS–placebo responding association is robust and largely independent of methodological quality features.

We next examined whether 24 binary variables pertaining to methodological features moderated correlation coefficients in the total sample of included studies (**Table 2**). Employing a direct verbal suggestion to induce placebo responses showed a weak, non-significant negative moderation effect. The use of a partially or entirely student sample was a weak positive moderator. There were no other significant moderating effects related to inclusion of a control condition, experimenter sex, inclusion of non-clinical sample, placebo administration type, mode of suggestion, REVS assessment features and target symptom domain. Bayesian moderation and subgroup ROPE analyses indicated that the student sample moderator showed limited posterior mass within the ROPE, suggesting a practically meaningful moderation effect. In contrast, moderation effects for condition counterbalancing and use of indirect suggestion showed posterior distributions concentrated within the ROPE, indicating negligible differences between subgroups, while the association between REVS and placebo responding within each subgroup remained practically meaningful (see **Supplementary Tables 2 G 3**). Cumulatively, most (34 of 38) of the moderation effects fell within the ROPE, suggesting evidence for a negligible moderation effect. Ultimately, these findings indicate that the REVS–placebo association was generally robust across methodological features, with some evidence that this association was larger in studies including student samples, condition counterbalancing, and indirect verbal suggestions.

### Sensitivity analyses

We conducted a series of pre-specified and exploratory sensitivity analyses to further clarify the magnitude of the association between REVS and placebo responses. We were able to conduct four of six pre-specified analyses based on the available data. In each case, analyses were restricted to methodologically rigorous studies, defined by adequate participant and experimenter blinding, sample sizes above the median (*n* > 40), and inclusion of a control condition (see **Supplementary Results**). For our exploratory analyses, we restricted our analyses to studies that included condition counterbalancing, sample type (i.e., student samples), and suggestion type (direct or indirect). In all but one case, the aggregate REVS– placebo correlation remained significant and within the 95% CIs of the main effect size estimate (range: 0.15 to 0.23). However, studies using only direct suggestions showed a weak but non-significant trend. Collectively, these results indicate that the REVS-placebo response correlation is relatively stable.

## Discussion

The present meta-analysis quantified the association between trait REVS (suggestibility) and placebo responses by synthesizing data from available empirical studies. These analyses demonstrate a reliable positive association, such that individuals higher in REVS tend to display larger placebo responses. Notably, the magnitude of this effect was relatively stable irrespective of different methodological features. These data build upon consistent evidence showing the clear efficacy of verbal suggestions in the induction of placebo effects [12, 13]. The observed pattern of results bridges a gap in the literature between stable trait patterns of REVS [22, 51, 52] and the use of verbal suggestion to effect placebo responses. These results provide novel meta-analytic evidence that an individual difference characteristic known to moderate responsiveness to verbal suggestions—a central feature of placebo inductions—is also associated with placebo responses.

The central finding of this meta-analysis is that REVS is a reliable predictor of placebo responses. This result warrants a nuanced reconsideration of the controversy regarding a putative good placebo responder [9]. The relative failure to previously identify reliable trait correlates of placebo responding [10, 11] is arguably due to a relative neglect of variables that are known to moderate responses to the component induction methods of placebo procedures (e.g., suggestion). The present findings nicely complement evidence pointing to REVS as a reliable predictor of response to various suggestion-based procedures [53, 25, 24, 54, 26] and suggests that the predictive efficacy of REVS generalises to multiple domains where suggestions are employed. It is notable that this effect was relatively stable across studies and mostly invariant to methodological variability. Indeed, 89% of the included REVS-placebo response correlations were positive in direction and heterogeneity was low. This consistency supports the interpretation that the observed association reflects a stable pattern across studies. Although placebo induction paradigms vary substantially, verbal suggestion constitutes a core component of most placebo procedures [2, 55, 56] and was formally identified, or suspected, to be a primary feature of the placebo procedure in all studies included in this analysis. Accordingly, REVS may capture individual differences in responsiveness to one of the primary mechanisms through which placebo responses are elicited. From this perspective, the present findings suggest that REVS predicts placebo responses not because it indexes a general placebo responder trait *per se*, but because it moderates responsiveness to a ubiquitous feature of placebo interventions. Nevertheless, the magnitude of this association was modest and thereby does not indicate that individuals who are high in REVS reflect the long-contested good placebo responder. Rather, the magnitude of this effect suggests that variability in REVS represents one important contributor to placebo responses among a broader constellation of psychological, genetic, and neurobiological factors that may aid in the prediction of placebo responses [57, 1, 58, 59]. Accordingly, the present results support a shift away from binary distinctions between placebo responder and non-responders toward a multivariate account of placebo response [58], in which individual differences in REVS constitute one of several factors contributing to variability in placebo outcomes.

An aim of this meta-analysis was to evaluate the boundary conditions of the association between REVS and placebo responses. Meta-regression analyses provided preliminary evidence that suggestion type may influence this association. In particular, placebo studies employing indirect suggestions tended to show larger REVS-placebo response correlations than those using direct suggestions. Direct and indirect suggestions are not uniform categories, and the source of this trend is unclear; however, a tendency for participants to overweight information regarding others may underlie this apparent difference [17]. An examination of the subset of studies that reported the exact wording of placebo suggestions (*n* = 8) pointed to variability in both suggestion types: indirect suggestions tended to emphasize attention [60], interpretation of ongoing sensations [31], and subjective appraisal of experience [28, 61] whereas direct suggestions tended to frame perceptual changes as the result of an external, efficacious intervention [62, 29, 63, 64], at times invoking evidence such as clinical use, scientific validation, or a mechanistic explanation [29, 64]. Interpretation of these differences is limited by the small number of studies reporting verbatim suggestions and the variability within each suggestion type. By contrast, we found no significant moderating effects related to the mode of suggestion (e.g., verbal, textual) but these results should be similarly interpreted cautiously given that most of the studies (*k* = 57; 86%) used verbal suggestions to induce placebo responses. Ultimately, these findings preliminarily indicate that the content of suggestions may be more relevant than their mode of delivery and that high REVS individuals may differentially respond to different types of suggestions in placebo studies.

Expectations are widely theorised to underlie placebo effects [65] and may represent a key pathway through which suggestions exert their influence on symptom perception [2].

Although we observed reliable placebo expectancy and placebo effects, neither variable moderated the association between REVS and placebo responses. Previous research suggests that individuals higher in REVS may form stronger placebo expectations [28, 29]. However, we found no evidence that placebo expectancy moderated the association between REVS and placebo responding at the study level [28]. This discrepancy does not preclude the possibility that expectancy mediates the relationship between REVS and placebo responses at the individual level [66, 29], a hypothesis that cannot be directly evaluated in the present meta-analysis due to reliance on summary level data published in the original studies [67]. As such, individuals higher in REVS may be more responsive to placebo suggestions, although the magnitude of this association is modest. From a predictive processing perspective [68], individuals with elevated REVS would be expected to form a more precise prior (i.e., belief) for symptom relief in response to verbal suggestions in the context of a placebo procedure; the prior is subsequently over-weighted relative to sensory input, leading to greater symptom reduction [69]. Although placebo expectancy did not moderate the REVS–placebo response association, there was a non-significant trend for this association to increase with the magnitude of the placebo effect. Collectively, these findings highlight the need to better understand the mechanisms underlying the REVS–placebo response association.

The association between REVS and placebo responses was relatively stable but differentially scaled with multiple methodological features. Across moderators, a consistent pattern emerged whereby features that reduced bias or measurement noise were associated with larger observed correlations. For example, studies that ensured proper blinding of experimenters to participants’ placebo responses when measuring REVS reported larger correlations than those that did not adequately blind experimenters. This reaffirms the importance of experimenter blinding to minimise the impact of experimenter effects on placebo responses and placebo-trait associations [70], especially given the predominance of subjective outcomes that may be sensitive to contextual and reporting influences (but see also [71]).

Turning to sample composition, studies conducted with student samples reported larger REVS-placebo response correlations than those conducted with non-students. Although this finding should be interpreted cautiously given the limited number of the latter studies (*k* = 2; 3%), it may reflect the greater influence of context-dependent factors (e.g., motivation) [72], which could attenuate the observable contribution of REVS and warrants further investigation in clinical populations where REVS is sometimes a less robust predictor of clinical outcomes following suggestion-based interventions (e.g., [73]). On the other hand, the negative moderating effects of clearly reporting sample recruitment site and strategy may imply that studies with more transparent reporting provide more conservative estimates of the REVS– placebo response association, whereas less well-reported studies may inadvertently obscure sources of selection bias, sampling bias, or other methodological factors associated with inflated effect size [74]. Taken together, these findings suggest that the association between REVS and placebo responses is robust across study characteristics and that methodological rigor appears to enhance the detectability of this association.

### Limitations

These results should be interpreted in light of methodological considerations pertaining to our analyses and limitations of the primary studies. With respect to meta-analytic methodological considerations, correlations based on change scores may underestimate the REVS–placebo response association due to greater measurement error [75], potentially contributing to variability in observed effects. In addition, our moderation analyses had lower statistical power than the overall effect analyses, warranting caution in interpreting the predominantly null moderation findings. In respect to the limitations of the primary studies, the majority (*n* = 18; 69%) did not report the precise wording of placebo suggestions, limiting our ability to reliably code suggestion type. Similarly, overall methodological quality was relatively low. Few studies ensured that experimenters were blinded to experimental conditions (*n* = 9; 36%) or to participants’ REVS scores when assessing placebo responses (*n* = 11; 44%), and none assessed the integrity of blinding procedures [70]. In addition, only a minority of studies (*n* = 7; 35%) included a control condition, with most relying on baseline comparisons (*n* = 12; 60%), precluding condition counterbalancing and increasing the risk of order effects and related confounds. Finally, most studies (*n* = 12; 48%) were only powered (*N* range: 44–93) to detect moderate-to-large effects, suggesting that many estimates of the REVS–placebo response association are probably imprecise. However, sensitivity analyses did not suggest that this association was attributable to the preponderance of studies with small sample sizes.

### Future directions

The aforementioned limitations highlight the need for rigorously designed and transparently reported studies assessing the apparent association between REVS and placebo responding. The reporting of verbatim suggestions and whether suggestions were standardised across sessions should become a minimum basic reporting standard. Future research should also include well-controlled comparison conditions, rigorous participant and experimenter blinding and assessment of blinding integrity. Beyond methodological improvements, the weak aggregate correlation between REVS and placebo responses suggests that REVS represents only one of several individual difference factors contributing to variability in placebo responses. Future research should therefore examine the joint and potentially interactive contributions of REVS alongside other traits, such as dissociative absorption [7, 76], while also accounting for state-dependent influences such as motivation [77]. Individuals high in REVS are a heterogeneous population [78, 79] and thus the consideration of REVS as part of a multivariate prediction model will be essential to progress research in this area. More broadly, these findings highlight the need to examine individual differences not only in response to verbal suggestion, but also other placebo induction methods, such as conditioning and social observation [11]. Finally, future research should examine how suggestion type shapes the magnitude, durability, and phenomenology of placebo responses and their association with REVS.

### Implications

The demonstrated role of REVS in predicting placebo responses has several important implications for both the empirical study of placebo responses and their occurrence in clinical contexts [80]. In empirical settings, integrating REVS into existing accounts of placebo responses [69, 4] could offer a more holistic understanding of the contribution of stable trait individual differences to variability in placebo responses. Further, measurement of REVS could aid in further delineating and identifying good placebo responders across demographic variables that covary with REVS, and clinical status. For example, youth [81] or clinical populations wherein elevated REVS is a hallmark feature (e.g., individuals with functional neurological disorder) may exhibit greater placebo responding [82, 83, 36, 84].

In clinical settings, from a precision medicine standpoint [85], identifying the role of REVS in predicting placebo responding may pave the way for more personalized treatment approaches, such as screening REVS during treatment allocation [5] and differentially leveraging suggestion to augment active treatment effects [86, 87].

## Conclusion

This meta-analysis identified a reliable association between trait REVS and placebo responses, highlighting the role of individual differences in shaping responsiveness to suggestion-based processes. Although the pooled effect size was small, its consistency across meta-regression and sensitivity analyses indicates that this relationship is robust to variation in study design and methodological features. Overall, these findings position REVS as a modest but consistent predictor of placebo responses, supporting a more integrative account in which stable individual differences and contextual factors, jointly shape symptom reduction in response to contextual cues.

## Data Availability

All data relevant to the present meta-analysis is provided in the article. Data are freely available from previous research studies.

## Acknowledgements

We’d like to thank Siobhan Murphy, Ben Smith, and Reena Swaroop for their assistance with screening and data extraction. ChatGPT (OpenAI) was used as a support tool during data analysis and manuscript preparation. The authors retain full responsibility for the integrity, interpretation, and reporting of the work.

## Author contributions

All authors conceived the project. MVS carried out the database searches; data coding by reviewers was completed with assistance from MVS, TT and DBT. MVS performed the meta-analysis in consultation with TT and DBT. MVS drafted the initial manuscript with comments provided by DBT. TT reviewed and approved the final version of the manuscript.

## Footnotes

1 Two studies that reported overlapping samples were clustered together for the analyses

