## Supplementary materials for "Trait responsiveness to verbal suggestions predicts placebo responses: A multi-level meta-analysis"

**Supplementary results**

A 15-item scale was developed to assess study methodological quality. Items were rated in a binary fashion (0 = criterion not met, 1 = criterion met) and computed into a percentage score. Items are based on Cochrane criteria and PRISMA recommendations (Stein et al., 2023; Thompson et al., 2019; Wieder et al., 2021).

1) Were relevant reliability (e.g., Cronbach's alpha, test-retest reliability) and validity data (e.g., construct validity) for the suggestibility measure reported in this study or cited from previous research?
2) Was a referenced or standardized symptom measure used?
3) Was there a clear specification of study objectives?
4) Was it clearly described where participants were drawn from (e.g., University, community bulletins, etc.)?
5) Was it clearly described how participants were recruited (e.g., advertisement, course credits, volunteers, etc.)?
6) Was there a clear description of the inclusion and exclusion criteria?
7) Was the procedure described in enough detail to allow for independent replication?
8) Was the experimenter blind to participants' placebo response when measuring their suggestibility?
9) Was the experimenter blind to participants' suggestibility when measuring their placebo response?
10) Were precautions implemented to ensure that participants were blind to experimental condition (control vs. placebo)?
11) Were precautions implemented to ensure the experimenter was blind to experimental condition (control vs. placebo)?
12) Were relevant participant characteristics adequately described (age, sex, etc.,)?
13) Were the groups comparable in terms of demographics? [Applicable only to between-groups studies].
14) Was the study pre-registered?
15) Are the data freely available on an open data repository?

**Supplementary Table 1.** *Consensus methodological quality ratings per study (n = 26)*

| Study | 1 | 2 | 3 | 4 | 5 | 6 | 7 | 8 | 9 | 10 | 11 | 12 | 13 | 14 | 15 | Total score (%) |
|---|---|---|---|---|---|---|---|---|---|---|---|---|---|---|---|---|
| Baker et al., 1993a | 0 | 1 | 1 | 1 | 1 | 0 | 1 | 1 | 1 | 1 | 1 | 1 | 1 | 0 | 0 | 73% |
| Bentler et al., 1963a | 0 | 0 | 1 | 1 | 0 | 0 | 0 | 0 | 0 | 0 | 0 | 0 | NA | 0 | 0 | 14% |
| Bush & Ditto, 1985 | 0 | 0 | 1 | 0 | 1 | 1 | 0 | 0 | 1 | 1 | 1 | 0 | NA | 0 | 0 | 42% |
| Corsi et al., 2017a | 0 | 1 | 1 | 0 | 0 | 1 | 1 | 1 | 0 | 1 | 0 | 1 | NA | 1 | 0 | 57% |
| De Pascalis & Scacchia, 2019a | 0 | 1 | 1 | 0 | 0 | 1 | 0 | 1 | 0 | 0 | 0 | 0 | NA | 0 | 0 | 28% |
| De Pascalis et al., 1999a | 0 | 1 | 1 | 0 | 0 | 0 | 0 | 1 | 0 | 0 | 0 | 0 | NA | 0 | 0 | 21% |
| De Pascalis et al., 2001 | 0 | 1 | 1 | 1 | 0 | 0 | 0 | 1 | 1 | 0 | 0 | 0 | NA | 0 | 0 | 35% |
| De Pascalis et al., 2002a | 0 | 1 | 1 | 1 | 0 | 1 | 1 | 1 | 0 | 1 | 1 | 0 | NA | 0 | 0 | 57% |
| De Pascalis et al., 2016a | 0 | 1 | 1 | 1 | 1 | 0 | 1 | 1 | 0 | 1 | 1 | 1 | NA | 0 | 0 | 64% |
| De Pascalis et al., 2021a | 0 | 1 | 1 | 0 | 0 | 1 | 0 | 1 | 0 | 1 | 0 | 1 | NA | 0 | 0 | 42% |
| Guestini & Parris, 2025a | 0 | 1 | 1 | 1 | 1 | 1 | 1 | 0 | 1 | 0 | 0 | 1 | NA | 0 | 0 | 57% |
| Huber et al., 2013 | 0 | 1 | 1 | 0 | 0 | 0 | 0 | 0 | 1 | 1 | 0 | 1 | NA | 0 | 0 | 35% |
| Leigh et al., 2003 | 0 | 1 | 1 | 1 | 1 | 1 | 0 | 1 | 1 | 1 | 0 | 1 | NA | 0 | 0 | 64% |
| Lifshitz et al., 2017a | 0 | 0 | 1 | 1 | 1 | 0 | 0 | 0 | 1 | 0 | 0 | 0 | NA | 0 | 0 | 28% |
| Lund et al., 2015a | 0 | 1 | 1 | 1 | 1 | 1 | 1 | 1 | 0 | 1 | 1 | 0 | NA | 0 | 0 | 64% |
| McGlashan et al., 1969 | 0 | 0 | 1 | 1 | 1 | 1 | 0 | 1 | 0 | 0 | 1 | 0 | NA | 0 | 0 | 42% |
| Parsons et al.,2021-Study 1 | 1 | 1 | 1 | 1 | 0 | 1 | 1 | 1 | 1 | 1 | 1 | 1 | NA | 0 | 1 | 85% |
| Parsons et al.,2021-Study 2 | 1 | 1 | 1 | 1 | 0 | 1 | 1 | 1 | 1 | 1 | 1 | 1 | NA | 0 | 1 | 85% |
| Ryan & Souheaver, 1976 | 0 | 1 | 1 | 1 | 1 | 1 | 0 | 1 | 0 | 1 | 0 | 0 | NA | 0 | 0 | 50% |
| Sharav et al., 2023a | 0 | 1 | 1 | 1 | 1 | 0 | 1 | 1 | 0 | 1 | 0 | 1 | NA | 0 | 0 | 57% |
| Sheiner et al., 2016a | 1 | 0 | 1 | 1 | 1 | 1 | 1 | 1 | 0 | 1 | 0 | 1 | NA | 0 | 0 | 64% |
| Spanos et al., 1988a | 0 | 0 | 1 | 1 | 1 | 0 | 0 | 0 | 1 | 0 | 1 | 0 | NA | 0 | 0 | 35% |
| Spanos et al., 1989-Study 1 | 0 | 1 | 1 | 1 | 1 | 0 | 0 | 1 | 0 | 0 | 0 | 0 | NA | 0 | 0 | 35% |
| Spanos et al., 1989-Study 2 | 0 | 1 | 1 | 1 | 1 | 0 | 0 | 1 | 0 | 0 | 0 | 0 | NA | 0 | 0 | 35% |
| Tasso et al., 2020 | 0 | 0 | 1 | 1 | 1 | 0 | 1 | 1 | 0 | 0 | 0 | 1 | NA | 0 | 0 | 42% |
| Woody et al., 1997a | 0 | 0 | 1 | 1 | 1 | 1 | 1 | 1 | 1 | 1 | 0 | 1 | NA | 0 | 0 | 64% |

***Notes.*** 0 = item not met; 1 = item met.

*Meta-analysis of placebo expectations*

A total of 15 studies assessed expectation of symptom reduction in response to the placebo (placebo expectancy), however only 10 studies ($k$ = 10) reported the relevant data to permit inclusion in a meta-analysis of placebo expectancy. As reported in the main text, this effect was statistically significant (**Supplementary Figure 1**). Egger's regression test showed significant funnel plot asymmetry ($Z$ = 10.52, $p$ <.001; **Supplementary Figure 2**), however a trim-and-fill analysis identified no missing studies, leaving the aggregate effect size unchanged. Finally, a Bayesian ROPE analysis indicated that the magnitude of the placebo expectancy effect was not practically equivalent to zero (posterior median $g$ = 0.96, credible intervals [0.04, 1.73]), with only 2.1% of the posterior mass falling within the ROPE. Cumulatively, these results suggest that the magnitude of the placebo expectancy effect was large but heterogenous and plausibly inflated by publication bias.

**Supplementary Figure 1.** *Forest plot of symptom expectancy Hedges' gs (k = 10).*

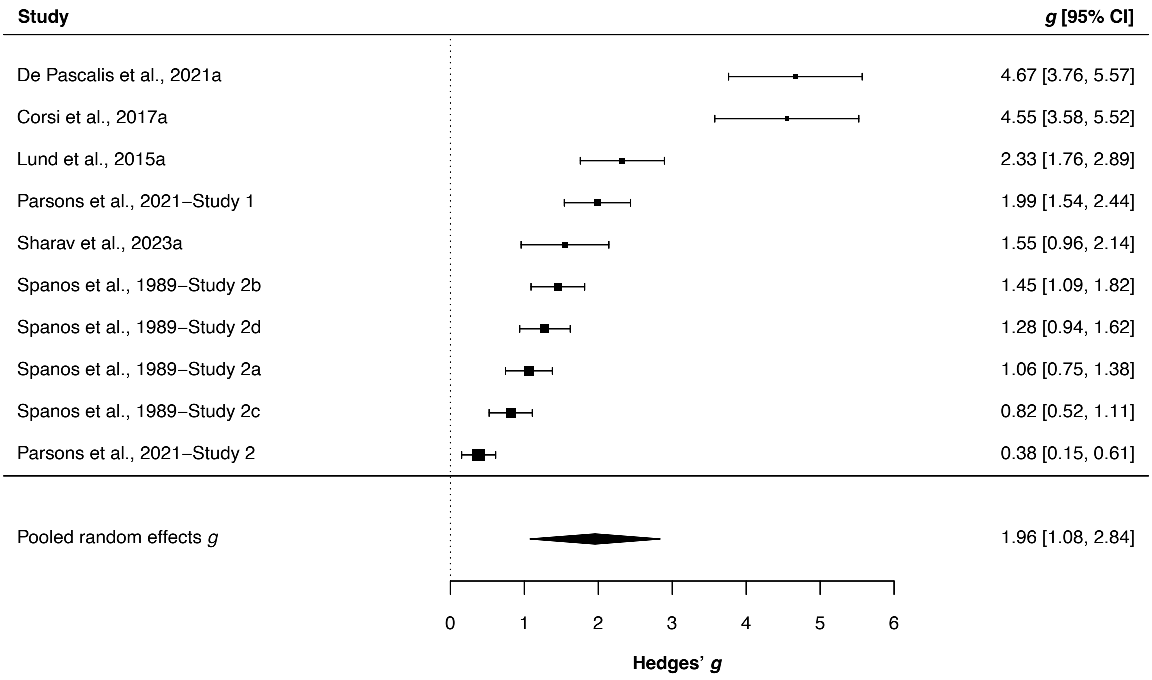


***Notes.*** Marker sizes reflect study weights, with smaller and larger markers denoting smaller and larger weights, respectively.

**Supplementary Figure 2.** *Funnel plot of symptom expectancy Hedges' gs as a function of standard errors (k = 10).*

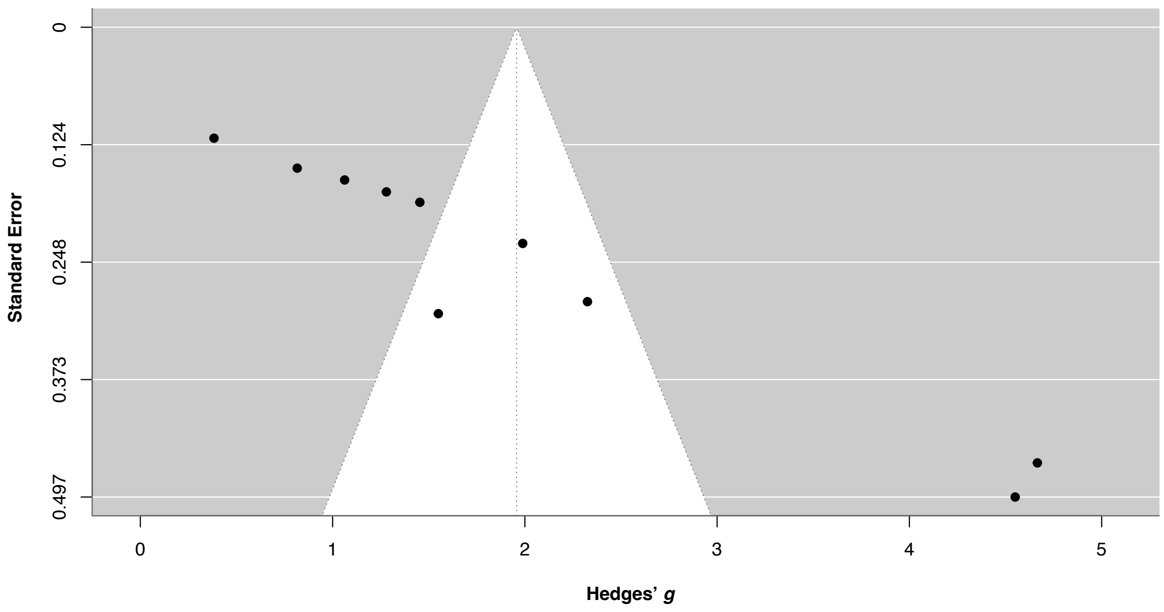


***Notes.*** Markers denote individual study effect sizes

*Meta-analysis of placebo effects*

Fifteen studies provided the necessary data to perform a meta-analysis on the placebo effect ($k$ = 15, $n$ = 9). As reported in the main text, the effect was statistically significant but moderate in magnitude (**Supplementary Figure 3**). Egger's regression test indicated no evidence of funnel plot asymmetry within a random-effects model ($Z$ = 0.56, $p$ = .57; **Supplementary Figure 4**) or multilevel model* of the data ($\beta$ = 1.12, $SE$ = 1.50, $Z$ = 0.75, $p$ = .45). However, a trim-and-fill analysis estimated two potentially missing studies, yielding a modestly attenuated but still significant pooled effect, $g$ = 0.29 [0.07, 0.51]. Finally, a Bayesian ROPE analysis indicated that the aggregate magnitude of the placebo effect was not practically equivalent to zero (posterior median $g$ = 0.35 [0.05, 0.64]), with only 4.5% of the posterior mass falling within the ROPE.

* A multilevel meta-analysis was used to synthesise the placebo effect data, but not the placebo expectancy data, because the latter violated the assumptions required for modelling multiple effect sizes from the same study.

Cumulatively, these results suggest that the magnitude of the placebo effect was moderate with no evidence of publication bias.

**Supplementary Figure 3.** *Multilevel forest plot of placebo effect (placebo-control) Hedges' gs (k = 15, n = 9).*

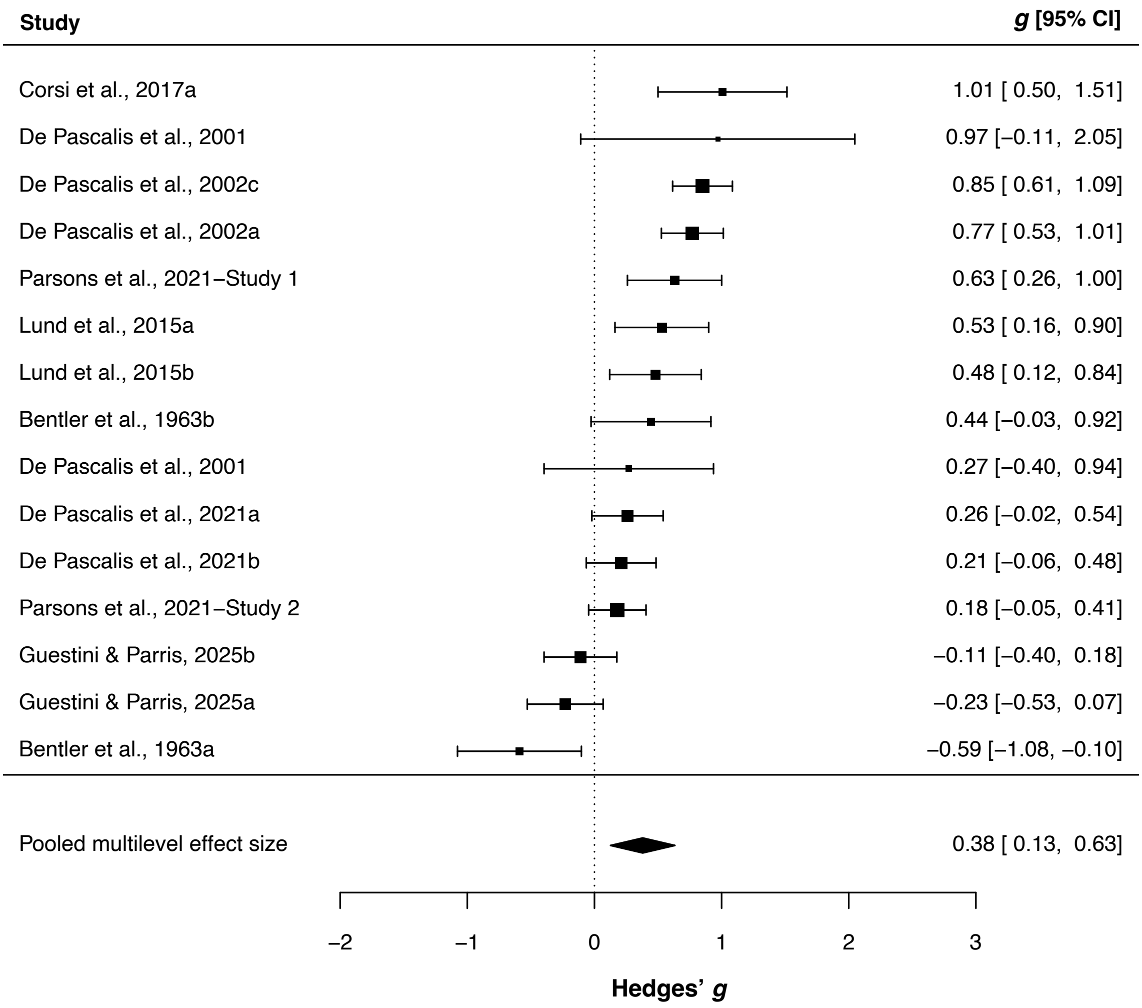


***Notes.*** Marker sizes reflect study weights, with smaller and larger markers denoting smaller and larger weights, respectively.

**Supplementary Figure 4**. *Random effects funnel plot of placebo effect (placebo-control) Hedges' gs as a function of standard errors (k = 15, n = 9).*

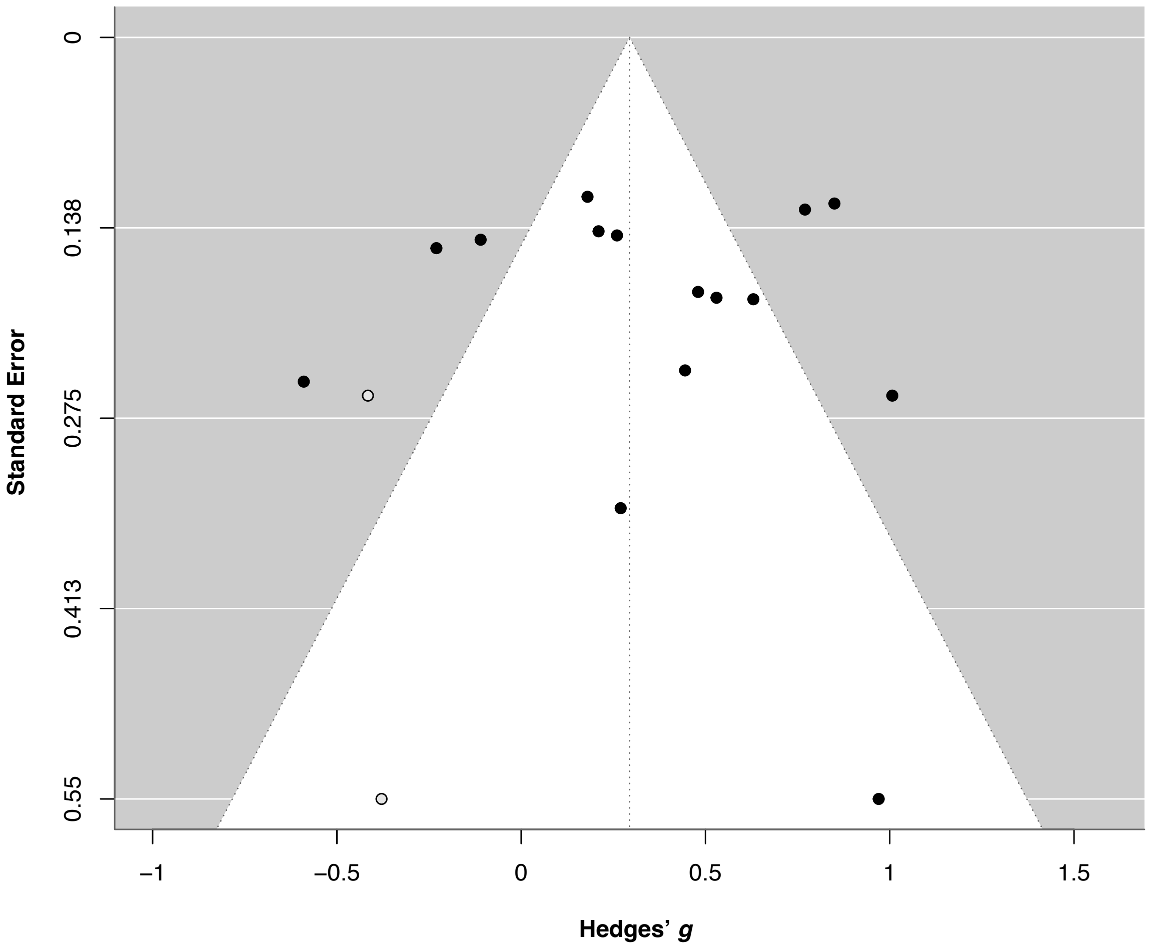


***Notes.*** Filled markers denote individual study effect sizes and empty markers denote estimated missing individual effect sizes due to publication bias imputed using the trim-and-fill method.

*Sensitivity analyses*

*Pre-specified analyses*

We pre-specified five sensitivity analyses based on methodological features expected to yield more robust effects (see https://tinyurl.com/5n84cuc4). We were able to conduct four of these analyses based on the available data. First, we restricted our meta-analysis (see main paper) to studies in which participants were adequately blinded to the experimental condition (e.g., placebo). This yielded a slightly larger effect size, corresponding to the upper bound of the confidence intervals from the main analysis ($k$ = 37, $n$ = 15), $r_{bt}$ = 0.23 [0.18, 0.28], Z = 8.48, $p$<.001, with minimal heterogeneity, $W\text{-}I^2$ = 1%, $B\text{-}I^2$ = 4%. Similarly, studies that ensured experimenters were adequately blinded to the experimental condition ($k$ = 20, $n$ = 9), yielded a similar effect size, $r_{bt}$ = 0.23 [0.13, 0.30], $Z$ = 4.95, $p$<.001, $W\text{-}I^2$ = 2%, $B\text{-}I^2$ = 14%. To incorporate these two variables, we restricted our analysis to studies that ensured adequate condition blinding in both participants and experimenters ($k$ = 17, $n$ = 7), which yielded an equivalent correlation, $r_{bt}$ = 0.23 [0.15, 0.31], $Z$ = 5.28, $p$<.001, $W\text{-}I^2$ = 2%, $B\text{-}I^2$ = 16%. Next, we restricted the analysis to studies with sample sizes above the median (*Med* = 40) of all included studies ($k$ = 33, $n$ = 12), $r_{bt}$ = 0.20 [0.15, 0.25], $Z$ = 7.40, $p$<.001, $W\text{-}I^2$ = 7%, $B\text{-}I^2$ = 11%. Our final pre-specified analysis was restricted to studies that incorporated a control condition ($k$ = 21, $n$ = 9) with little change in the aggregate correlation, $r_{bt}$ = 0.24 [0.16, 0.33], $Z$ = 5.48, $p$<.001, $W\text{-}I^2$ = 7%, $B\text{-}I^2$ = 26%. These analyses indicate that the correlation between REVS and placebo responding remains stable when restricted to the most methodologically robust studies and thus may be slightly larger than that reported in the main text.

*Exploratory analyses*

The foregoing analyses were complemented by a set of exploratory analyses in which the meta-analysis was re-performed on subsets of the data in order to clarify whether the observed REVS-placebo responding correlation was specific to particular samples or methodological features. Firstly, studies that used a student sample demonstrated a comparable effect size to the main analysis ($k$ = 44, $n$ = 17), $r_{bt}$ = 0.17 [0.11, 0.22], $Z$ = 5.80, $p$<.001, $W\text{-}I^2$ = 1%, $B\text{-}I^2$ = 10%. We observed a similar effect size when restricting studies to those that included non-clinical samples ($k$ = 59, $n$ = 21), $r_{bt}$ = 0.19 [0.14, 0.24], $Z$ = 7.19, $p$<.001, $W\text{-}I^2$ = 2%, $B\text{-}I^2$ = 12%. We could not restrict analyses to non-student ($k$ = 2, $n$ = 2) or clinical samples ($k$ = 3, $n$ = 3) due to insufficient sample sizes. Finally, when considering suggestion type, the aggregate effect size of studies that used direct suggestions ($k$ = 8, $n$ = 5) was weak and showed a non-significant trend, $r_{bt}$ = 0.13 [-0.00, 0.26], $Z$ = 1.92, $p$ = .054, $W\text{-}I^2$ = 3%, $B\text{-}I^2$ = 6%, whereas studies that used indirect suggestions ($k$ = 12, $n$ = 4) showed an aggregate effect size that was slightly larger than the effect size of the main dataset, $r_{bt}$ = 0.26 [0.16, 0.36], $Z$ = 4.90, $p$<.001, $W\text{-}I^2$ = 2%, $B\text{-}I^2$ = 1%. Cumulatively, these results suggest that when restricted to particular samples or methodological features, the effect size was similar to that of the main analysis, albeit slightly higher in studies that used indirect suggestions.

*Equivalence testing of moderation and subgroup analyses*

We performed a series of Bayesian equivalence tests of both moderation analyses (**Supplementary Table 2**) and subgroup analyses (**Supplementary Table 3**; see main paper) in order to evaluate the practical relevance of the observed moderator effects, drawing inferences from the proportion of the posterior mass falling within the predefined region of practical equivalence (ROPE). Among the former, the effects indicated that differences between subgroup levels were generally small and often practically negligible. Although frequentist meta-regression analyses identified significant correlation differences, primarily associated with selected methodological quality indicators (e.g., recruitment clarity, blinding procedures) and sample characteristics (e.g., student samples), Bayesian ROPE analyses suggested that, with the exception of student samples, the differences between subgroup levels lay within the region of practical equivalence. Subgroup equivalence analyses further corroborated this pattern: the absolute subgroup effects were uniformly small and fell within the predefined ROPE, indicating that statistically significant moderation effects corresponded to negligible practical differences. Taken together, these findings suggest that while certain study features are associated with relative variation in effect size, the absolute magnitude of the REVS–placebo association is modest and largely robust across methodological quality and features.

### **Supplementary Table 2.** *Equivalence test (ROPE) results for moderation analyses*

| Moderator | *Posterior median $r_{bt}$* [95% CrI] | *N (k)* | Posterior in ROPE (%) |
|---|---|---|---|
| **Methodological quality** | | | |
| Item 1: relevant reliability data provided for REVS measure | 0.02 [-0.16, 0.19] | 25 (66) | 73% |
| Item 2: standardized symptom outcome | 0.04 [-0.09, 0.17] | 25 (66) | 80% |
| Item 4: clear sample recruitment site | -0.10 [-0.21, 0.00] | 25 (66) | 47% |
| Item 5: clear participant recruitment strategy | -0.09 [-0.18, 0.00] | 25 (66) | 57% |
| Item 6: clear inclusion & exclusion criteria | 0.10 [0.01, 0.20] | 25 (66) | 48% |
| Item 7: clear protocol reporting | 0.04 [-0.07, 0.14] | 25 (66) | 89% |
| Item 8: experimenter blinded to placebo response for REVS | 0.16 [0.02, 0.30] | 25 (66) | 20% |
| Item 9: experimenter blind to REVS when measuring placebo | -0.10 [-0.21, 0.01] | 25 (66) | 52% |
| Item 10: participant condition blinding | 0.08 [-0.02, 0.17] | 25 (66) | 68% |
| Item 11: experimenter condition blinding | -0.03 [-0.16, 0.10] | 25 (66) | 85% |
| Item 12: adequate description of sample demographics | -0.02 [-0.13, 0.08] | 25 (66) | 93% |
| Total methodological quality score* | 0.01 [-0.10, 0.12] | 25 (66) | 93% |
| **Methodological features** | | | |
| Inclusion of control condition | 0.08 [-0.04, 0.21] | 25 (66) | 66% |
| Female experimenter | -0.06 [-0.32, 0.21] | 6 (15) | 55% |
| Female & male experimenters | 0.12 [-0.13, 0.39] | 6 (15) | 40% |
| Non-clinical sample | 0.14 [-0.16, 0.45] | 25 (62) | 34% |
| Some/all student sample | 0.38 [-0.03, 0.72] | 25 (62) | 8% |
| Magnitude of the placebo effect* | 0.21 [-0.02, 0.45] | 9 (15) | 18% |
| Measured expectancy | 0.05 [-0.06, 0.16] | 25 (66) | 80% |
| Magnitude of the expectancy effect* | 0.09 [-0.24, 0.48] | 7 (10) | 40% |
| **Placebo administration type** | | | |
| Pill as placebo administration type | -0.07 [-0.24, 0.11] | 25 (66) | 62% |
| Sham device as placebo administration type | -0.04 [-0.27, 0.18] | 25 (66) | 58% |
| Cream as placebo administration type | 0.07 [-0.04, 0.19] | 25 (66) | 69% |
| Inhalant as placebo administration type | -0.14 [-0.42, 0.05] | 25 (66) | 34% |
| Use of conditioning procedure | 0.00 [-0.15, 0.14] | 25 (66) | 84% |
| **Suggestion mode and type** | | | |
| Textual suggestion | -0.07 [-0.21, 0.08] | 25 (66) | 64% |
| Verbal suggestion | -0.06 [-0.24, 0.11] | 25 (66) | 66% |
| Nonverbal suggestion | -0.05 [-0.19, 0.10] | 25 (66) | 76% |
| Use of direct verbal suggestion | -0.13 [-0.30, 0.06] | 9 (20) | 46% |
| **REVS assessment features** | | | |
| Group administration of REVS scale | 0.02 [-0.11, 0.16] | 21 (58) | 85% |
| Use of two REVS scales | -0.12 [-0.40, 0.17] | 25 (66) | 39% |
| Inclusion of an induction on REVS scale | -0.06 [-0.17, 0.07] | 25 (66) | 76% |
| Assessment of REVS in the same context as placebo administration | 0.01 [-0.14, 0.16] | 23 (63) | 83% |
| Counterbalancing of conditions | 0.09 [-0.01, 0.20] | 25 (66) | 55% |
| **Target symptom domain** | | | |
| Pain as symptom outcome | 0.09 [-0.04, 0.23] | 25 (66) | 57% |
| Cognition as symptom outcome | 0.00 [-0.19, 0.19] | 25 (66) | 72% |
| Affect as symptom outcome | 0.03 [-0.08, 0.14] | 25 (66) | 89% |

***Notes.*** *CrI* = Bayesian credible interval; *N* = total sample size across included studies; *k* = number of studies. *ROPE* denotes the predefined region of practical equivalence around zero. Boldface indicates contrasts that did not meet the Bayesian equivalence criterion (i.e., only a small proportion of the posterior distribution fell within the ROPE), consistent with a non-negligible effect. * For continuous moderators, $\Delta r$ represents the difference in predicted pooled effect size between studies 1 SD above versus 1 SD below the moderator (after standardization within the analysed subset).

**Supplementary Table 3.** *Equivalence test (ROPE) results for absolute subgroup pooled effects*

| Subgroup feature | Feature absent | Feature present |
|---|---|---|
| | N (*k*), Posterior median $r_{bt}$ [95% CrI], Posterior in ROPE (%) | |
| **Methodological quality** | | |
| Item 1: relevant reliability data provided for REVS measure | 22 (59), 0.18 [0.10, 0.24], 2% | 3 (7), 0.19 [-0.21, 0.47], 20% |
| Item 2: standardized symptom outcome | 8 (19), 0.15 [0.01, 0.27], 21% | 17 (47), 0.20 [0.11, 0.26], 2% |
| Item 4: clear sample recruitment site | 6 (17), 0.27 [0.14, 0.37], 1% | 20 (49), 0.16 [0.08, 0.23], 6% |
| Item 5: clear participant recruitment strategy | 10 (27), 0.24 [0.14, 0.32], 1% | 16 (39), 0.15 [0.07, 0.22], 8% |
| Item 6: clear inclusion & exclusion criteria | 12 (34), 0.14 [0.06, 0.21], 13% | 14 (32), 0.24 [0.12, 0.32], 2% |
| Item 7: clear protocol reporting | 14 (32), 0.14 [0.01, 0.25], 22% | 12 (34), 0.21 [0.14, 0.28], 0% |
| Item 8: experimenter blind to placebo response when measuring REVS | 6 (13), 0.05 [-0.13, 0.23], 67% | 19 (53), 0.21 [0.15, 0.27], 0% |
| Item 9: experimenter blind to REVS when measuring placebo | 14 (43), 0.23 [0.15, 0.29], 0% | 11 (23), 0.11 [-0.04, 0.23], 42% |
| Item 10: participant condition blinding | 11 (29), 0.13 [0.02, 0.22], 25% | 15 (37), 0.23 [0.15, 0.30], 0% |
| Item 11: experimenter condition blinding | 12 (29), 0.24 [0.15, 0.31], 0% | 5 (11), 0.21 [0.02, 0.38], 8% |
| Item 12: adequate description of sample demographics | 13 (34), 0.19 [0.08, 0.28], 4% | 13 (32), 0.19 [0.10, 0.26], 3% |
| **Methodological features** | | |
| Inclusion of control condition | 17 (45), 0.16 [0.05, 0.23], 12% | 9 (21), 0.22 [0.09, 0.33], 3% |
| Female experimenter | 3 (8), 0.16 [-0.24, 0.42], 24% | 3 (7), 0.10 [-0.26, 0.39], 40% |
| Female & male experimenters | 4 (9), 0.08 [-0.18, 0.28], 53% | 2 (6), 0.20 [-0.31, 0.53], 16% |
| Non-clinical sample | - | 21 (59), 0.20 [0.13, 0.25], 0% |
| Some/all student sample | - | 17 (44), 0.17 [0.10, 0.23], 2% |
| Measured expectancy | 12 (22), 0.13 [-0.04, 0.26], 34% | 14 (44), 0.21 [0.14, 0.27], 0% |
| **Placebo administration type** | | |
| Pill as placebo administration type | 21 (53), 0.19 [0.12, 0.25], 1% | 4 (13), 0.12 [-0.15, 0.35], 38% |
| Sham device as placebo administration type | 21 (61), 0.19 [0.12, 0.24], 1% | 4 (5), 0.15 [-0.28, 0.52], 29% |
| Cream as placebo administration type | 15 (36), 0.15 [0.04, 0.23], 17% | 10 (30), 0.23 [0.13, 0.30], 1% |
| Inhalant as placebo administration type | 23 (61), 0.20 [0.14, 0.25], 0% | 2 (5), -0.12 [-0.68, 0.53], 23% |
| Use of conditioning procedure | 20 (54), 0.18 [0.10, 0.25], 3% | 5 (12), 0.18 [-0.01, 0.34], 15% |
| **Suggestion mode and type** | | |
| Textual suggestion | 20 (54), 0.20 [0.12, 0.26], 1% | 5 (12), 0.13 [-0.12, 0.36], 35% |
| Verbal suggestion | 3 (9), 0.22 [-0.18, 0.51], 14% | 22 (57), 0.18 [0.10, 0.24], 2% |
| Nonverbal suggestion | 20 (51), 0.20 [0.11, 0.26], 1% | 5 (15), 0.14 [-0.06, 0.30], 28% |
| Use of direct verbal suggestion | - | 5 (8), 0.13 [-0.12, 0.35], 35% |
| Use of indirect verbal suggestion | - | 4 (12), 0.26 [0.03, 0.42], 5% |
| **REVS assessment features** | | |
| Group administration of REVS scale | 12 (33), 0.18 [0.02, 0.28], 13% | 9 (25), 0.20 [0.10, 0.30], 2% |
| Use of two REVS scales | 23 (63), 0.19 [0.12, 0.24], 1% | - |
| Inclusion of an induction on REVS scale | 7 (16), 0.17 [-0.15, 0.37], 26% | 18 (50), 0.17 [0.11, 0.23], 2% |
| Assessment of REVS in the same context as placebo administration | 19 (54), 0.19 [0.10, 0.25], 2% | 4 (9), 0.19 [-0.04, 0.37], 14% |
| **Target symptom domain** | | |
| Pain as symptom outcome | 7 (17), 0.08 [-0.22, 0.31], 47% | 18 (49), 0.21 [0.14, 0.26], 0% |
| Cognition as symptom outcome | 23 (63), 0.19 [0.11, 0.24], 1% | - |
| Affect as symptom outcome | 21 (47), 0.18 [0.09, 0.24], 3% | 8 (19), 0.23 [0.10, 0.36], 3% |

***Notes.*** Reported values correspond to posterior median pooled correlations ($r_{bt}$) and 95% credible intervals (CrI) estimated within subgroups defined by each binary characteristic (coded 1). ROPE percentages indicate the posterior probability that the estimated effect lies within a region of practical equivalence ($|r| \leq .10$). All subgroup estimates describe absolute effects and do not represent contrasts between moderator levels.

### List of included papers

Baker, S. L., & Kirsch, I. (1993). Hypnotic and placebo analgesia: Order effects and the placebo label. *Contemporary Hypnosis*, *10*(3), 117–126.

Bentler, P. M., O'Harra, J. W., & Krasner, L. (1963). Hypnosis and placebo. *Psychological Reports*, *12*(1), 153–154. https://doi.org/10.2466/pr0.1963.12.1.153

Bush, C., Ditto, B., & Feuerstein, M. (1985). A controlled evaluation of paraspinal EMG biofeedback in the treatment of chronic low back pain. *Health Psychology*, *4*(4), 307–321. https://doi.org/10.1037/0278-6133.4.4.307

Corsi, N., & Colloca, L. (2017). Placebo and nocebo effects: The advantage of measuring expectations and psychological factors. *Frontiers in Psychology*, *8*, 308. https://doi.org/10.3389/fpsyg.2017.00308

De Pascalis, V., Chiaradia, C., & Carotenuto, E. (2002). The contribution of suggestibility and expectation to the placebo analgesia phenomenon in an experimental setting. *Pain*, *96*(3), 393–402. https://doi.org/10.1016/S0304-3959(01)00485-7

De Pascalis, V., Magurano, M. R., & Bellusci, A. (1999). Pain perception, somatosensory event-related potentials, and skin conductance responses to painful stimuli in high, mid, and low hypnotizable subjects: Effects of differential pain reduction strategies. *Pain*, *83*(3), 499–508. https://doi.org/10.1016/S0304-3959(99)00157-8

De Pascalis, V., Magurano, M. R., Bellusci, A., & Chen, A. C. (2001). Somatosensory event-related potentials and autonomic activity during varying pain-reduction cognitive strategies in hypnosis. *Clinical Neurophysiology*, *112*(8), 1475–1485. https://doi.org/10.1016/S1388-2457(01)00586-7

De Pascalis, V., & Scacchia, P. (2016). Hypnotizability and placebo analgesia in waking and hypnosis as modulators of auditory startle responses in healthy women: An ERP study. *PLoS One*, *11*(8), e0159135. https://doi.org/10.1371/journal.pone.0159135

De Pascalis, V., & Scacchia, P. (2019). The influence of reward sensitivity, heart rate dynamics, and EEG-delta activity on placebo analgesia. *Behavioural Brain Research*, *359*, 320–332. https://doi.org/10.1016/j.bbr.2018.11.014

De Pascalis, V., Scacchia, P., & Vecchio, A. (2021). Influences of hypnotic suggestibility, contextual factors, and EEG alpha on placebo analgesia. *American Journal of Clinical Hypnosis*, *63*(4), 302–328. https://doi.org/10.1080/00029157.2020.1863182

Guestini, A., & Parris, B. A. (2025). The relationship between dissociated measures of inhibition and hypnotic and placebo analgesia effects. *Biopsychosocial Science and Medicine*, *87*(7), 470–482. https://doi.org/10.1097/PSY.0000000000001405

Huber, A., Lui, F., & Porro, C. A. (2013). Hypnotic susceptibility modulates brain activity related to experimental placebo analgesia. *Pain*, *154*(9), 1509–1518. https://doi.org/10.1016/j.pain.2013.03.031

Leigh, R., MacQueen, G., Tougas, G., Hargreave, F. E., & Bienenstock, J. (2003). Change in forced expiratory volume in 1 second after sham bronchoconstrictor in suggestible but not suggestion-resistant asthmatic subjects: A pilot study. *Psychosomatic Medicine*, *65*(5), 791–795. https://doi.org/10.1097/01.psy.0000079454.48714.1b

Lifshitz, M., Sheiner, E. O., Olson, J. A., Thériault, R., & Raz, A. (2017). On suggestibility and placebo: A follow-up study. *American Journal of Clinical Hypnosis*, *59*(4), 385–392. https://doi.org/10.1080/00029157.2016.1225252

Lund, K., Petersen, G. L., Erlandsen, M., De Pascalis, V., Vase, L., Jensen, T. S., & Finnerup, N. B. (2015). The magnitude of placebo analgesia effects depends on how they are conceptualized. *Journal of Psychosomatic Research*, *79*(6), 663–668. https://doi.org/10.1016/j.jpsychores.2015.05.002

McGlashan, T. H., Evans, F. J., & Orne, M. T. (1969). The nature of hypnotic analgesia and placebo response to experimental pain. *Psychosomatic Medicine*, *31*(3), 227–246. https://doi.org/10.1097/00006842-196905000-00003

Parsons, R. D., Bergmann, S., Wiech, K., & Terhune, D. B. (2021). Direct verbal suggestibility as a predictor of placebo hypoalgesia responsiveness. *Psychosomatic Medicine*, *83*(9), 1041–1049. https://doi.org/10.1097/PSY.0000000000000977

Ryan, J. J., & Souheaver, G. T. (1976). Effects of transcrerebral electrotherapy (electrosleep) on state anxiety according to suggestibility levels. *Biological Psychiatry*, *11*(2), 233–237.

Sharav, Y., Haviv, Y., & Tal, M. (2023). Placebo or nocebo interventions as affected by hypnotic susceptibility. *Applied Sciences*, *13*(2), 931. https://doi.org/10.3390/app13020931

Sheiner, E. O., Lifshitz, M., & Raz, A. (2016). Placebo response correlates with hypnotic suggestibility. *Psychology of Consciousness: Theory, Research, and Practice*, *3*(2), 146–153. https://doi.org/10.1037/cns0000074

Spanos, N. P., Perlini, A. H., & Robertson, L. A. (1989). Hypnosis, suggestion, and placebo in the reduction of experimental pain. *Journal of Abnormal Psychology*, *98*(3), 285–293. https://doi.org/10.1037/0021-843X.98.3.285

Spanos, N. P., Stenstrom, R. J., & Johnston, J. C. (1988). Hypnosis, placebo, and suggestion in the treatment of warts. *Psychosomatic Medicine*, *50*(3), 245–260. https://doi.org/10.1097/00006842-198805000-00003

Tasso, A. F., Pérez, N. A., Moore, M., Griffo, R., & Nash, M. R. (2020). Hypnotic responsiveness and nonhypnotic suggestibility: Disparate, similar, or the same? *International Journal of Clinical and Experimental Hypnosis*, *68*(1), 38–67. https://doi.org/10.1080/00207144.2020.1685330

Woody, E. Z., Drugovic, M., & Oakman, J. M. (1997). A reexamination of the role of nonhypnotic suggestibility in hypnotic responding. *Journal of Personality and Social Psychology*, *72*(2), 399–407. https://doi.org/10.1037/0022-3514.72.2.399
